# Facemasks prevent influenza-like illness: implications for COVID-19

**DOI:** 10.1101/2020.05.07.20094912

**Authors:** Jie Wei, Michael Doherty, Monica S.M. Persson, Aliya Sarmanova, Subhashisa Swain, Changfu Kuo, Chao Zeng, Guanghua Lei, Weiya Zhang

**Affiliations:** Health Management Center, Xiangya Hospital, Central South University, Changsha, Hunan, China; Academic Rheumatology Department, Division of Rheumatology, Orthopaedics and Dermatology, School of Medicine, University of Nottingham, Nottingham, UK; Centre of Evidence Based Dermatology, Division of Rheumatology, Orthopaedics and Dermatology, School of Medicine, University of Nottingham, Nottingham, UK; Musculoskeletal Research Unit, Bristol Medical School (THS), University of Bristol, Bristol, UK; Division of Rheumatology, Allergy and Immunology, Chang Gung Memorial Hospital, Taoyuan, Taiwan; Department of Orthopaedics, Xiangya Hospital, Central South University, Changsha, Hunan, China

## Abstract

The coronavirus disease 2019 (COVID-19) pandemic is causing a huge toll on individuals, families, communities and societies across the world. Currently, whether wearing facemasks in public should be a measure to prevent transmission of severe acute respiratory syndrome coronavirus-2 (SARS-CoV-2) remains contraversial.^1^ This is largely because there have been no randomized controlled trials (RCTs) for coronavirus to directly support this. However, lessons may be taken from published RCTs examining influenza-like illness (ILI).^2,3^ Recent studies suggested that SARS-CoV-2 shares similar transmission route with influenza virus,^4^ and the incidence of community transmission of SARS-CoV-2 in individuals with ILI is high.^5^ Therefore, we undertook this meta-analysis of RCTs examining the efficacy of wearing facemasks to prevent ILI in community settings, irrespective of confirmatory testing for the causative virus.

We undertook a systematic literature search for RCTs related to facemasks and ILI between 1966 and April 2020 using PUBMED, EMBASE, and Cochrane library. RCTs undertaken in community (not hospital) settings comparing wearing and not wearing facemasks for ILI were included. Incidence of ILI (e.g., fever, cough, headache, sore throat, aches or pains in muscles or joints) was estimated per group. Relative risk (RR) and 95% confidence interval (CI) were calculated.

We screened 899 related abstracts and eventually included 8 RCTs (**Figure S1**). Basic characteristics and quality of included RCTs are listed in **Supplement**. Participants wearing facemasks had a significantly lower risk of developing ILI than those not wearing facemasks (pooled RR=0.81, 95% CI: 0.70–0.95) and there was no heterogeneity (**Figure 1**). The decreased risk of ILI was more pronounced if everyone wore facemask irrespective of whether they were infected or not (RR=0.77, 95% CI: 0.65-0.91), compared to those wearing facemasks when infected (RR=0.95, 95% CI: 0.58-1.56) or uninfected (RR=1.26, 95% CI: 0.69-2.31).

This study shows that wearing facemasks, irrespective of infection status, is effective in preventing ILI spread in the community. This situation mirrors what is happening now in public settings where we do not know who has been infected and who has not. Although there are no RCTs of facemasks for SARS-CoV-2, as with other simple measures such as social distancing and handwashing, these data support the recommendation to wear facemasks in public to further reduce transmission of SARS-CoV-2 and flatten the curve of this pandemic, especially when social distancing is impractical, such as shopping, or travelling with public transport for work that cannot be done from home.

---

**Figure 1.**
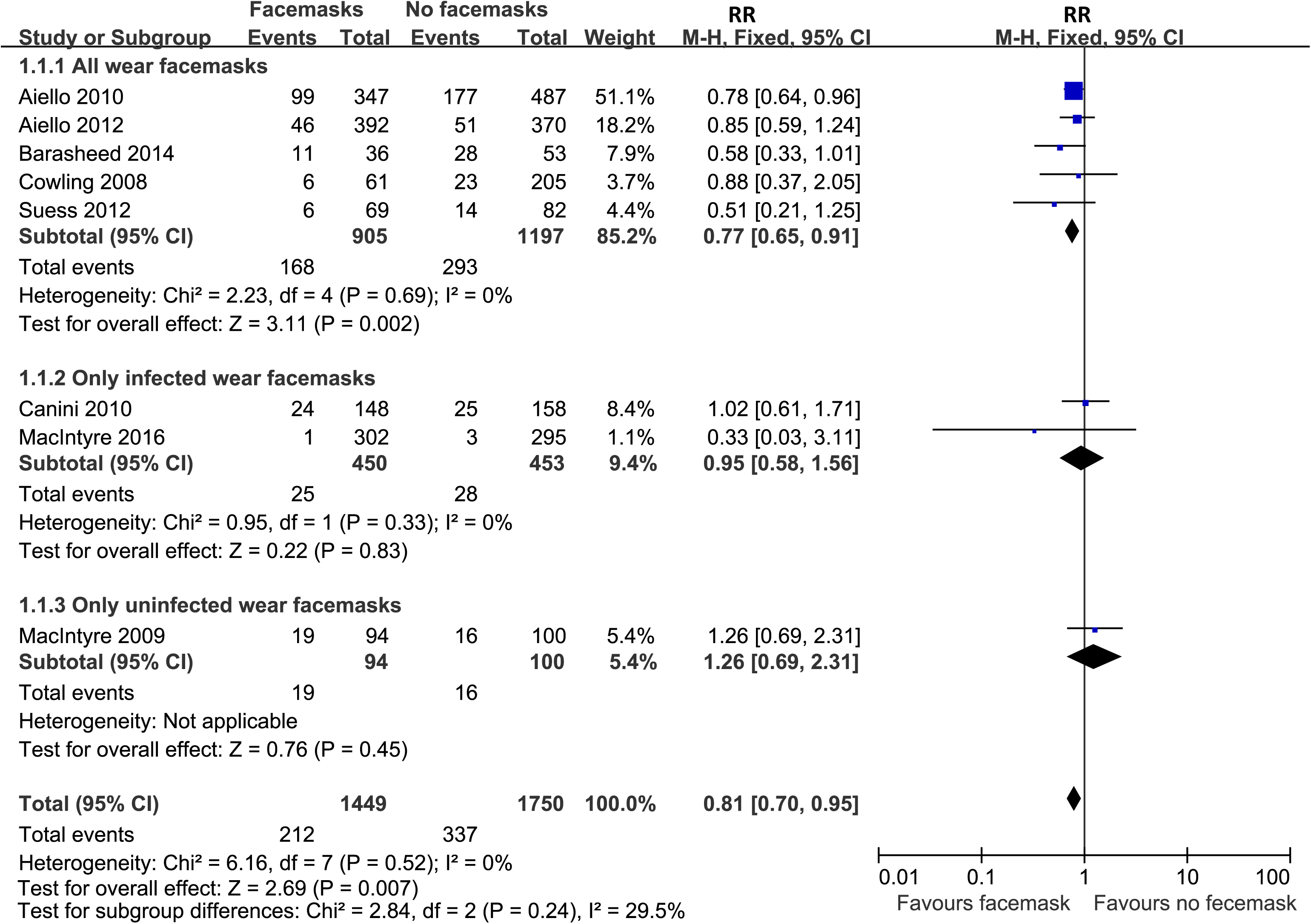
Relative risk (RR) and 95% confidence interval (CI) of developing influenza-like symptoms between wearing versus not wearing facemasks in general population.

## Data Availability

Data are avaliable by request

We declare no competing interests.

